# A phase 2 open-label clinical trial to determine the effect of Famciclovir on Epstein-Barr virus activity as measured by EBV shedding in the saliva of patients with Multiple Sclerosis

**DOI:** 10.1101/2023.08.18.23294265

**Authors:** Ruth Dobson, David Holden, Nicola Vickaryous, Jonathan Bestwick, Katila George, Tatiana Sayali, Lucia Bianchi, Mohammad Wafa, Julian Gold, Gavin Giovannoni

**Author notes:** Corresponding author: Dr Ruth Dobson, Preventive Neurology Unit, Wolfson Institute of Population Health Charterhouse Square, London EC1M 6BQ.

## Abstract

**Background:** There is increasing evidence that Epstein-Barr virus (EBV) plays a causal role in MS. No treatments have been shown to reduce EBV turnover. We studied the effect of famciclovir on salivary EBV shedding in people with MS (NCT05283551).

**Methods:** People with MS receiving natalizumab provided weekly saliva samples for 12 weeks before starting Famciclovir 500mg bd. 12 saliva samples were provided on treatment and 12 following treatment. A real-time quantitative PCR Taqman assay targeted to a non-repeated sequence of the EBV polymerase gene was used to detect EBV DNA in saliva. The proportion of saliva samples containing EBV DNA was compared using the Friedman test.

**Results:** 30 patients were recruited (19F; mean age 41 years; median EDSS 3.5). 29 patients received famciclovir, 24 completed the 12-week course. 21 participants provided at least one usable saliva sample in all 3 epochs. 10/21 participants had shedding in at least one sample pre-drug; 7/21 when taking famciclovir (not significant). No difference in EBV DNA copy number was seen. There were no drug-related serious adverse events.

**Conclusions:** No significant effect of famciclovir on EBV shedding was seen. Salivary EBV shedding in this natalizumab-treated cohort was lower than in previous studies; this requires replication.

## Background

There is increasing evidence that Epstein-Barr virus (EBV) plays a key role as an aetiological, or triggering, factor in multiple sclerosis (MS) development.^1,2^ EBV is a B-cell lymphotropic virus that infects more than 90% of the human population. Developing MS without evidence of prior EBV infection is rare.^3–5^ Recent large nested case-control studies demonstrate that people who are EBV-negative and subsequently develop MS always seroconvert to becoming EBV-positive before MS onset.^6^

Following primary infection, EBV establishes a lifelong presence in memory B-cells. It is not known how EBV triggers or contributes to MS development and/or ongoing disease activity. One theory is that people with MS appear to show evidence of dysregulated EBV control. People with MS have higher titres and broader antibody responses to both latent^7^ and lytic^8^ EBV proteins. In addition to elevated antibody titres against the EBNA complex and EBNA-1 antigen,^7,9^ people with MS have more EBNA-1 reactive CD4+ T-cells,^10^ which respond to a large repertoire of epitopes across the EBNA-1 protein. People with MS are more likely than people without MS to produce spontaneous EBV-associated lymphoblastoid cell lines (LCLs) when their peripheral lymphocytes are cultured ex vivo,^11–13^ potentially indicating significant viral dysregulation. However, these observations may indicate immune dysregulation in the context of MS rather than viral dysregulation.^14^

Salivary shedding of EBV occurs following acute symptomatic infection (infectious mononucleosis/ glandular fever).^15^ Salivary shedding also occurs periodically outside of the context of acute EBV infection, and is an important means of horizontal and vertical transmission.^16^ People with MS, including children, shed EBV in the saliva more frequently than people without MS.^17,18^ We have previously demonstrated that it is possible to demonstrate both EBV viral presence (detectable viral DNA) and periodic viral shedding (viral DNA copy number >5.8 copies/ul) in the saliva of people with MS.^17^

Whilst epidemiological, serological and immunological data indicate an important role for EBV in MS development, we do not know whether targeting EBV can influence the natural history of MS. Small randomised clinical trials using the anti-herpes nucleotide analogue acyclovir and its prodrug valacyclovir, showed some possible efficacy in patients with highly active MS, but overall results were inconclusive.^19,20^ Subtherapeutic drug levels, particularly in the central nervous system, could have affected the outcome of these studies. In the absence of a drug with proven anti-EBV activity, investigating the effect of inhibiting lytic EBV activity is impossible.

Famciclovir (FCV) is a prodrug of the nucleotide analogue penciclovir, which has enhanced CNS bioavailability and an intracellular half-life 10 times that of acyclovir. Famciclovir is rapidly converted to penciclovir, which is phosphorylated to the active triphosphate form by virus-expressed thymidine kinase, present only in virus-infected cells in the lytic phase. There are two case reports of the use of famciclovir in acute severe infectious mononucleosis.^19,21^ Famciclovir is widely used for prophylaxis of herpes virus infections in both immunocompetent and immunocompromised populations.

In order to understand the feasibility of a clinical trial to evaluate the effect of a potential anti-EBV treatment on clinical outcomes in MS, the impact of any proposed treatment on EBV turnover needs to first be evaluated. Given the relationship between EBV infection and MS, an MS cohort provides the ideal population in which to study this. Emerging evidence suggests that some MS disease modifying therapies have antiviral activity, either directly,^22,23^ or via targeting of lymphocyte compartments associated with EBV latency.^24,25^ We therefore set out to study the effect of famciclovir on salivary EBV shedding in a cohort of people with MS treated with natalizumab, a highly effective disease modifying therapy with no known antiviral activity.

## Methods

### Participants

30 patients with a confirmed diagnosis of relapsing remitting MS treated with natalizumab were recruited from the neurology department at the Royal London Hospital, Barts Health NHS Trust. All participants were aged over 18, had normal renal function as defined by eGFR, were not pregnant, breastfeeding or planning pregnancy, not on any antiviral medication, and had not received steroids in the three months prior to study entry. All participants provided written informed consent prior to study entry.

### Trial design and sample collection

This open label pre-post study lasted 36 weeks, with three phases. Sample and visit schedule is summarised in figure 1. All participants had a 12-week pre-treatment phase, prior to initiating treatment with famciclovir 500mg bd for 12 weeks. This was followed by a 12-week post-treatment phase (figure 1). All participants continued treatment with natalizumab throughout the study under usual neurological care. Where participants switched disease modifying therapy for clinical reasons, they were withdrawn from the study.

**Figure 1:**
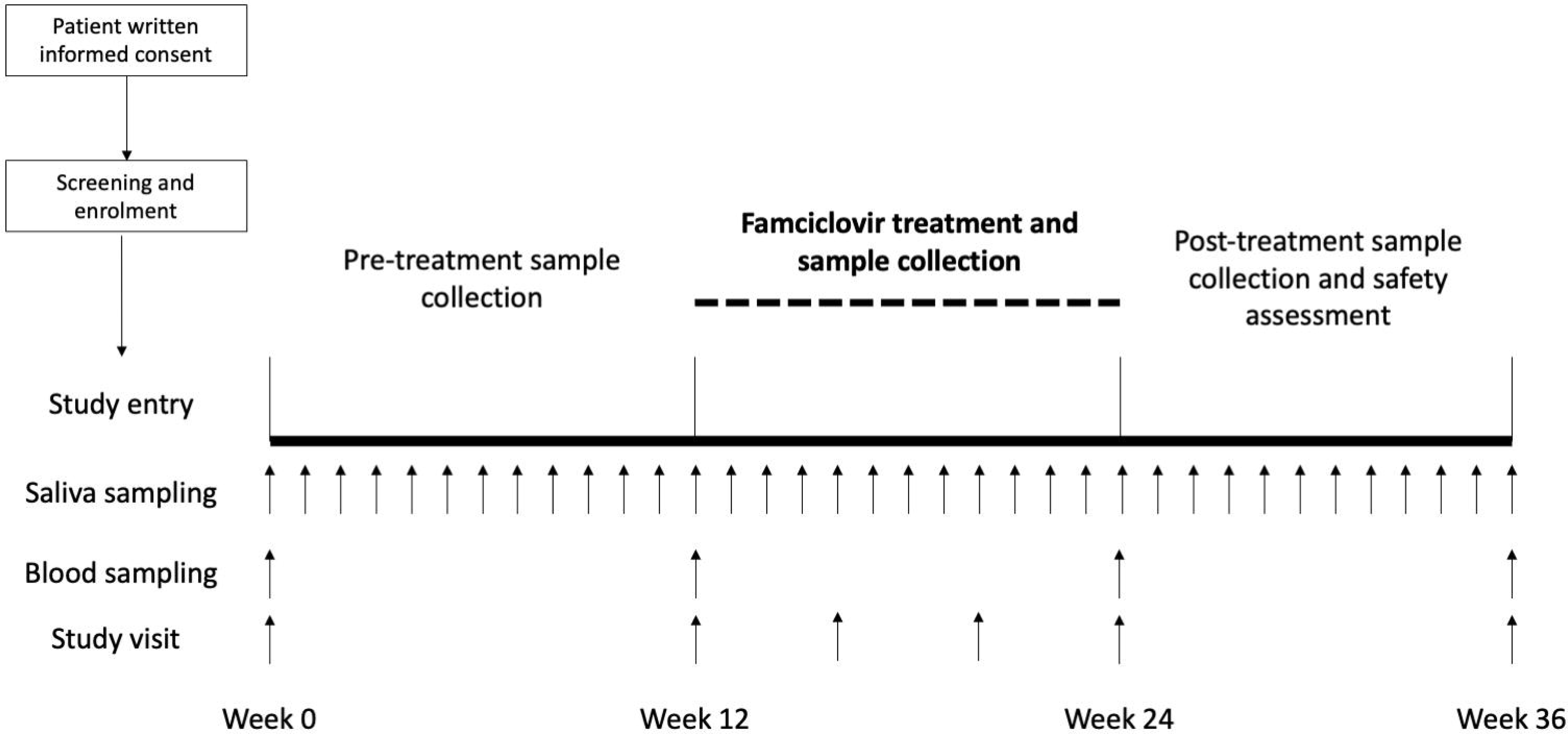
Study design and visit schedule.

Participants were asked to collect saliva samples on a weekly basis using salivettes throughout the study (figure 1). They were instructed to collect their saliva at the same time every week (ideally in the morning) due to significant diurnal variation in EBV shedding with a relative peak in the morning. No processing was required immediately post-sampling, and participants were instructed to store the samples in their freezer with double layered packaging, and to return them to the study site in person at study visits. Participants were provided with temperature monitoring devices in order to ensure sample integrity, and were provided with instructions in the event of freezer failure. Previous work has demonstrated the stability of saliva samples for DNA sequencing when collected and stored at home with repeated freeze-thaw cycles.^17^

### Visit schedule

Participants underwent scheduled study visits at the time of their natalizumab infusions, with additional telephone visits during the treatment period. Given the introduction of extended interval dosing for natalizumab, the drug monitoring visits could be in person or via telephone. Venous blood samples were taken at 4 timepoints during the study.

### Medication

Famciclovir 500mg tablets were provided to all participants, with instructions to take twice daily. Compliance was assessed using package return and self-report.

### Laboratory assessments

Analysis of EBV viral load in saliva was performed using a validated real-time quantitative PCR.^17^ EBV DNA was detected using a Taqman assay targeted to a non-repeated sequence of the EBV polymerase gene, together with a reference gene known to be present in genomic DNA. Samples were all run in duplicate, and a standard curve generated on each PCR plate using a dilution series derived from quantitated EBV DNA. This analysis has previously been fully reproducible (with coefficient of variation <1.4) for EBV levels above 3 copies/ml. An absolute copy number of EBV per sample (copies per µl of saliva) was then calculated directly from the standard curve.

Viral shedding was defined as a salivary EBV concentration greater than 5.8 copies/ml, with viral presence defined as any detectable EBV DNA in saliva.^17^

EBNA1 and VCA IgG titres in serum were measured using commercially available quantitative ELISA (Abnova, Taiwan). All serum samples had not been subject to previous freeze-thaw cycles. Briefly, samples were diluted in dilution buffer 1:500 and plated in duplicate along with the manufacturers standards and run according to manufacturer’s protocol. Results were expressed as arbitrary units per ml (AU/ml). Plate validity was assessed, and inter-assay coefficient of variation (CoV) was less than 5%.

### Sample size calculations

As this is a pilot, proof of concept study, formal sample size calculations were difficult to perform. It was estimated that 30 participants would allow us to detect a 50% reduction in the number of EBV-shedding positive samples on treatment across a range of pre-treatment shedding rates.

### Statistical analysis

Each saliva sample was assigned as showing evidence of EBV shedding, EBV viral DNA without evidence of shedding, or no detectable viral DNA. Missing samples were not imputed. The primary outcome measure, the rate of EBV shedding in saliva, was assessed as a proportion in the pre-treatment and the on-treatment stages. As the on-treatment group were expected to be skewed, the (paired) proportions were compared using the Wilcoxon signed rank Test.

A secondary analysis was performed using EBV DNA levels in saliva (treating EBV DNA levels as a continuous variable) in order to assess for any effect falling short of complete suppression. Median pre-treatment levels were compared to median on-treatment and post-treatment levels using a Friedman test. Finally, pre-treatment EBV DNA copy number/ml (weeks 0 and 12) was compared to immediate post-treatment (week 24) and delayed post-treatment (week 36) levels using a generalized linear mixed effects model (GLMM) to take account of temporal autocorrelation at the patient level.

Pre-treatment IgG titres (from blood samples taken at 0 and 12 weeks), immediate post-treatment titres (24 weeks), and titres after the post-drug study period (36 weeks) were normalised with log transformation and then compared using two-way ANOVA.

### Ethics

This study had ethical approval from the Westminster REC (reference 19/LO/1026). All participants provided informed consent to take part in the study.

## Results

### Participant demographics

31 potential participants entered screening, of whom 30 provided informed consent to take part in the study between November 2020 - January 2022. 29 participants received at least one dose of famciclovir, of whom 24 completed 12 weeks of treatment. 21 participants provided at least one usable saliva sample in all three epochs (pre-, during- and post-drug) and were included in the final analysis (figure 2). Participant demographics are summarised in table 1.

**Figure 2:**
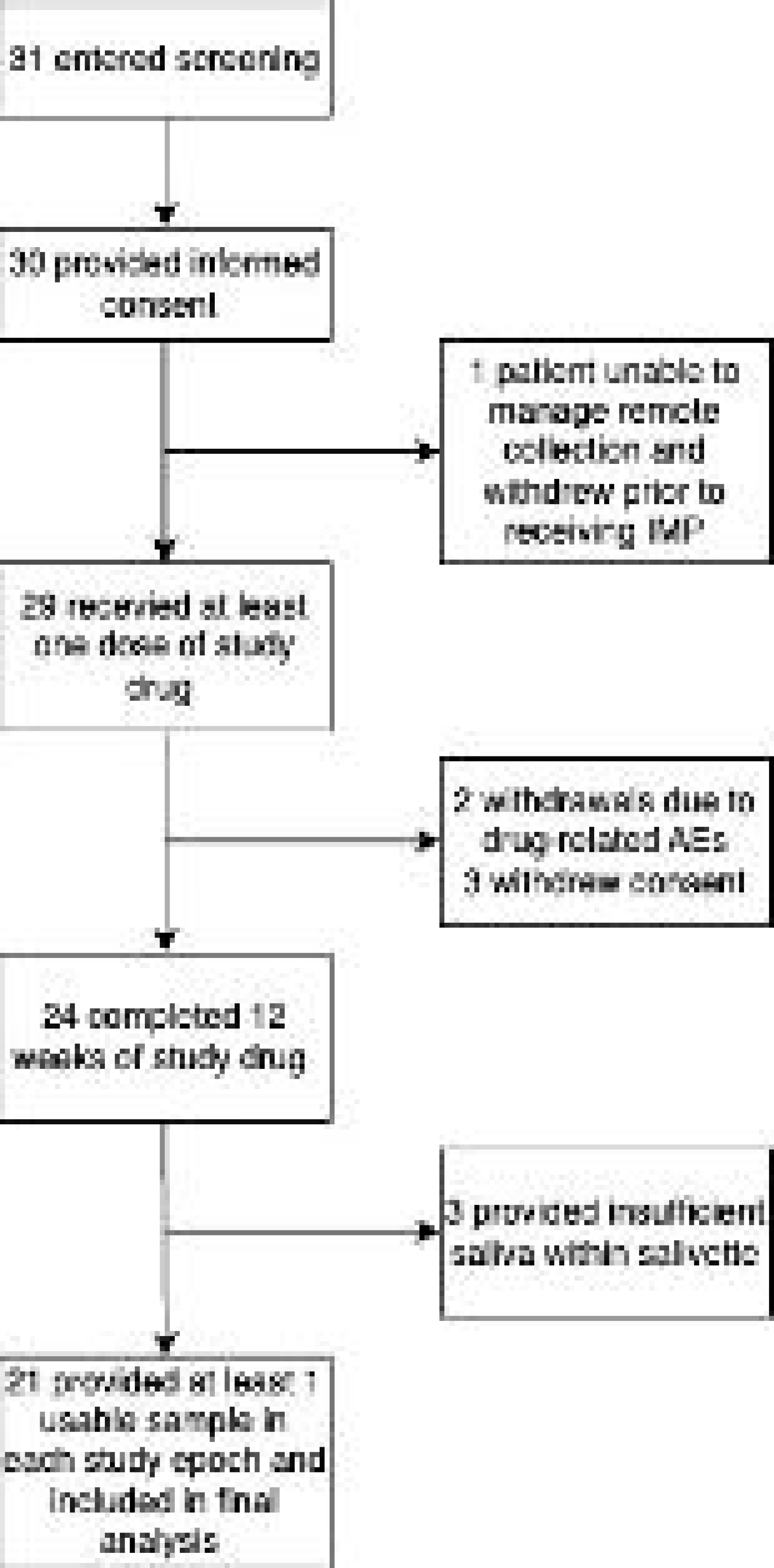
Participant flow through the study

**Table 1:** Participant demographics.

|  | Entire cohort (n=30) | Cohort with saliva samples available for analysis (n=21) |
| --- | --- | --- |
| Age at consent (median, range) | 45.8 (19.9 - 67.9) | 45.8 (19.9 - 65.0) |
| Sex (M:F; %F) | 11:19 (63%) | 7:14 (67%) |
| Ethnicity (white, minority ethnic; % White) | 25, 5 (83%) | 17, 4 (81%) |
| EDSS (median, range) | 3.5 (1.0 - 6.5) | 3.75 (1.0 - 6.5) |

Of the five participants who withdrew prior to completion of the treatment course (12 weeks of famciclovir), two had adverse events leading to study discontinuation (gastrointestinal symptoms and headaches). No serious adverse events related to the study drug were reported. All five participants chose to withdraw from the study and did not provide any further samples for analysis.

### Salivary shedding of EBV

All participants with at least one usable saliva sample showed evidence of detectable EBV DNA in saliva on at least one occasion. 10/21 (48%) participants had evidence of EBV shedding on at least one occasion pre-drug, 7/21 (33.3%) during and 5/21 (23.8%) post-drug (not significant; NS) (figure 3). When restricting to those participants with evidence of viral shedding in at least one sample, a median of 1/12 samples pre-treatment, 2/12 samples on treatment and 2.5/12 samples post-treatment showed evidence of viral shedding. Overall, 8.8% (16/181) pre-treatment samples showed evidence of viral shedding, 8.2% (15/183) whilst on treatment and 7.6% (14/184) in the post-treatment period.

**Figure 3:**
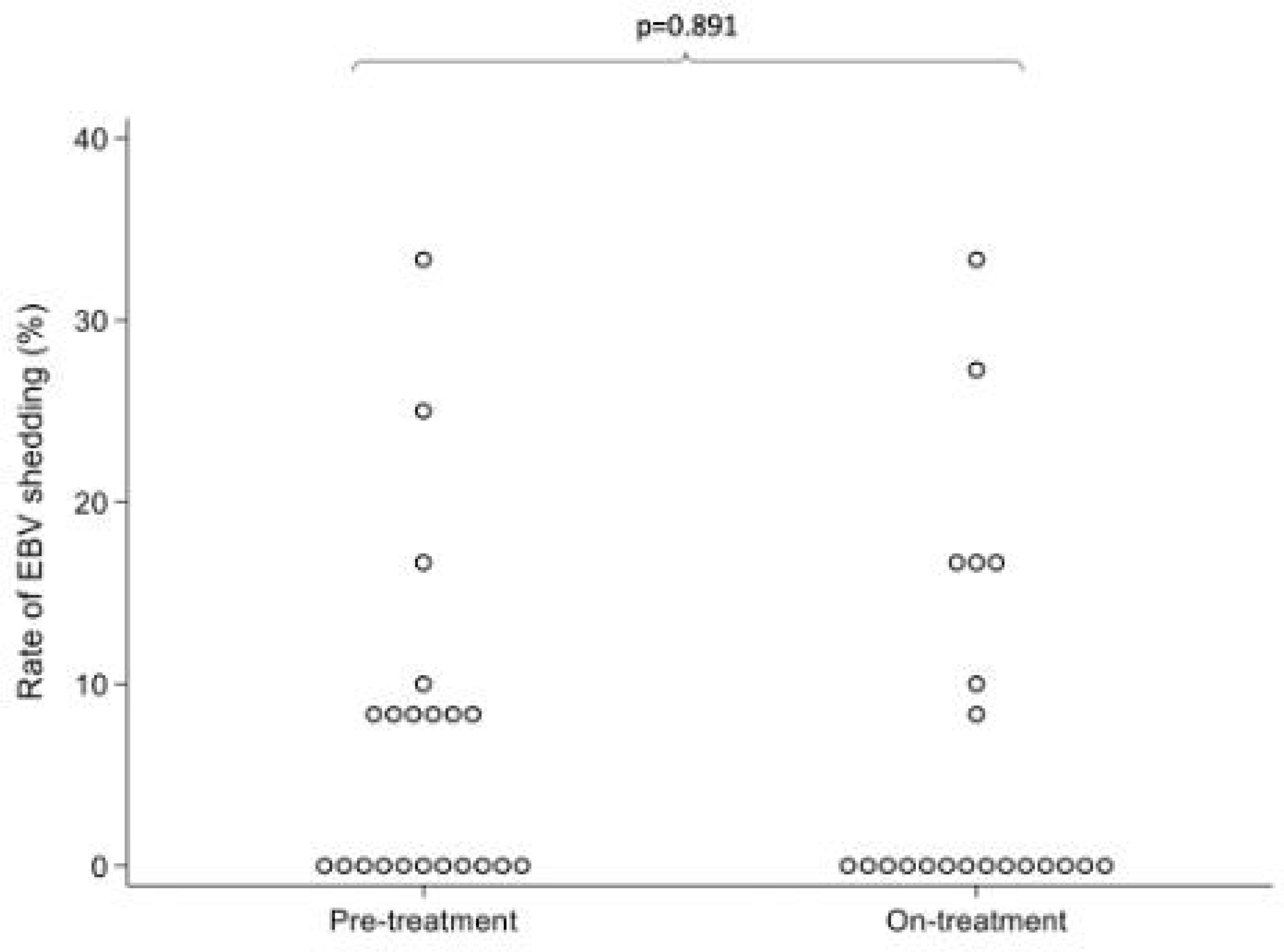
Median rate of EBV shedding (defined as >5.8 copies/ml) in the pre-treatment and on-treatment stages

EBV DNA was detected in 86/181 samples prior to treatment, 74/183 samples taken during famciclovir, and 100/184 samples taken after treatment. A median of 0 samples per participant had evidence of EBV shedding in all epochs (NS). There was no significant difference in the median EBV DNA copy number/ml between pre-treatment, on-treatment and post-treatment stages (figure 4). A GLMM showed no change in EBV DNA copy number/ml through the study (data not shown).

**Figure 4:**
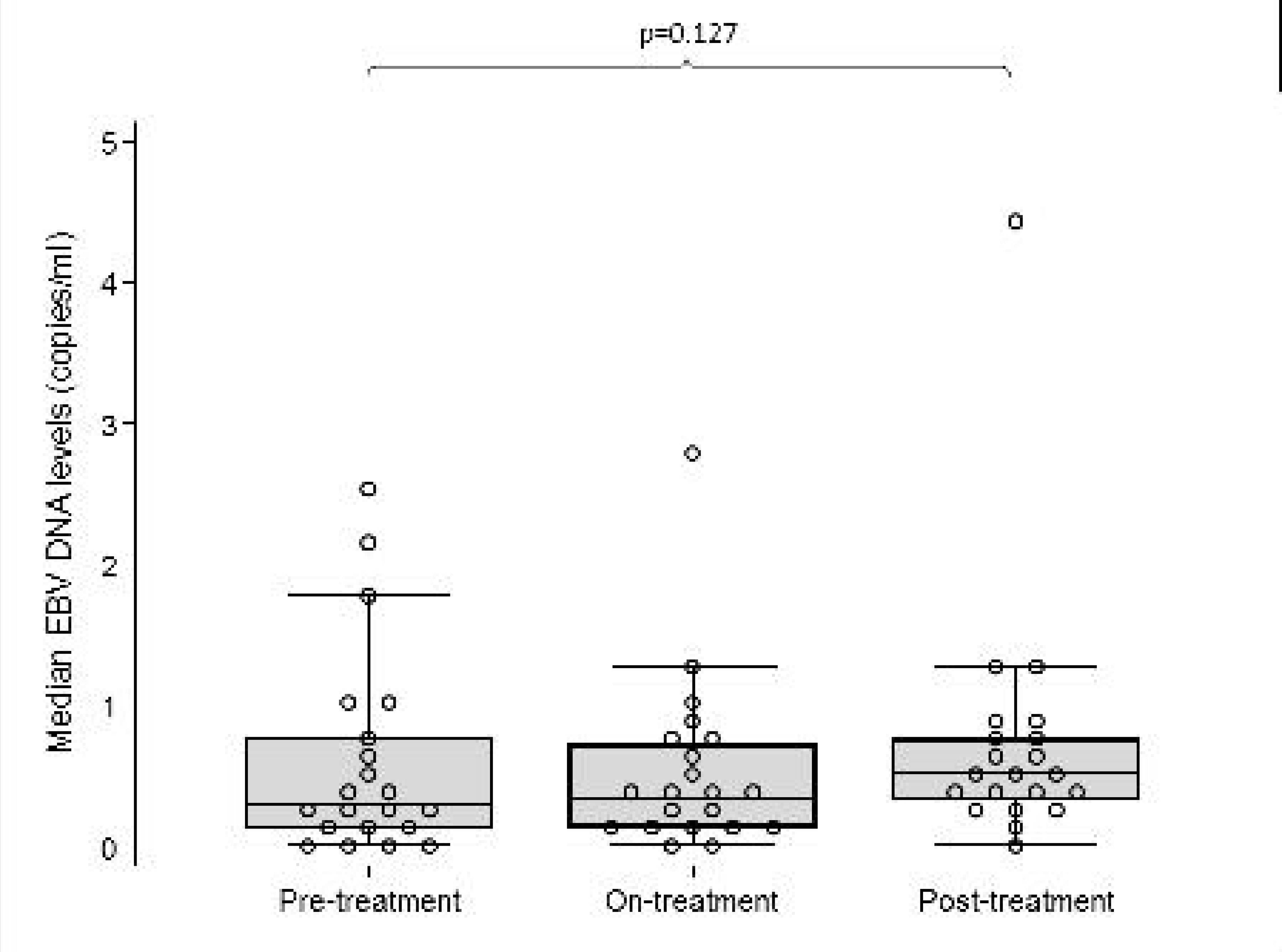
Combined dot and box and whisker plot demonstrating median EBV DNA levels (copies/ml) in the pre-treatment, on-treatment and post-treatment stages. Dots represent each datapoint; the box the interquartile range transected by the median, and whiskers the 5^th^ and 9^th^ percentiles.

### Serological response to EBV

No significant differences were seen between pre- and post-treatment IgG titres against either EBNA-1 or VCA in serum (data not shown).

## Discussion

Whilst famciclovir did not meet our primary endpoint of reducing the frequency of viral shedding in our cohort, the significant effect on both detectable viral DNA and overall copy number of EBV DNA detectable in saliva hint at an antiviral effect of the drug. Interventions to target existing EBV infection, or reduce viral load during acute EBV infection (infectious mononucleosis) remain an important unmet need in terms of potentially reducing the risk of MS in those exposed, or impacting on MS disease activity in people with MS. Without the development and testing of potentially effective anti-EBV interventions, understanding the role that EBV plays in MS development and disease activity remains challenging to study. It remains a possibility that the dose used, 500mg twice daily, was insufficient to inhibit EBV turnover. This dose was selected as it is the licensed prophylactic dose to prevent viral replication in people with HSV1, however 500mg four times daily is used for the treatment of active disease.

It must be noted that the overall level of salivary EBV shedding in the pre-treatment cohort (10/21 participants, with 8.8% samples showing evidence of shedding) is substantially lower than that demonstrated in other studies. We have previously demonstrated that 20-25% samples would be expected to show evidence of EBV shedding;^17^ similar work has demonstrated 17.5% samples from people living with HIV and 10-20% in healthy controls.^26^ The reason(s) for the unexpectedly low rates of shedding in our study are not immediately clear; our laboratory techniques were based on those previously used, and the high rates of low copy number DNA detection in samples indicates that we were detecting DNA when present. Natalizumab is not known to have any anti-viral action; indeed it is associated with an increased risk of PML secondary to JV virus infection,^27^ and additional case reports highlight potential morbidity from other viral reactivation syndromes in immune privileged sites.^28^ Of note, teriflunomide, an MS DMT with a mechanism of action in keeping with anti-viral efficacy, has shown significant effect on EBV shedding in saliva,^22^ and it may be that natalizumab has some anti-EBV properties, however we cannot further typify these at this stage.

The lack of any detectable effect on anti-EBV IgG titres, either against EBNA1 or VCA, is interesting. It remains unclear as to whether this reflects a lack of anti-EBV efficacy of famciclovir, or alternatively that the higher titres observed in people in MS are representative of aberrant immune responses rather than reflective of increased viral turnover in this group.

We had a higher than anticipated dropout rate in this study, although most participants completed the study and provided samples throughout. Two participants were unable to complete the course of famciclovir due to adverse events, both of which were clearly and commonly described in the summary of product characteristics, resolved on stopping the study medication and neither were serious. One participant was unable to continue due to switching DMT to a medication with potential anti-viral action. The remainder of the reasons for early study discontinuation related to challenges around sample collection and storage.

One of the major challenges with this study was the use of remotely collected samples. Whilst this reduced the need for in person visits, which was commended by patients, particularly in the context of the COVID pandemic, the volume of saliva collected remotely was less than expected for some patients. Despite the provision of clear instruction leaflets, demonstration of how to take samples at each visit and relatively simple sample collection techniques, sample volume remained an analytic issue. Furthermore, a number of samples were not returned, and some patients reported that they were unable to continue in the study due to concerns around sample collection and storage. This highlights the need to consider patient populations carefully during study design and recruitment, as unexpected challenges may become apparent.

Our study therefore provides an important proof of principle - that examining for EBV shedding in saliva is feasible in the context of a clinical trial. It highlights several challenges with the approach around recruitment and retention, sample collection, and overall rates of EBV shedding. We also provide evidence supporting increased immune responses rather than primary dysregulated infection as a potential link between EBV and increased IgG responses in people with MS. Our study will inform both power calculations and study design in future studies of anti-EBV agents in multiple sclerosis.

## Data availability

Anonymised participant-level data is available from the authors on request

## Author contributions

GG and JG conceived the idea of the study. RD drafted the protocol with input from GG, LB, KG and DH and led on study design and acted as study CI. RD, KG, LB, and MW carried out all clinical aspects of the study. DH and NV led laboratory analysis; JB led statistical analysis. RD drafted the initial manuscript, with important contributions from all co-authors.

## Potential conflicts of interest

RD: has received payments to her institution, including grants from Biogen, Merck, Celgene, National MS Society, MS Society UK, Horne Family Trust, and the BMA Foundation; honoraria from Biogen, Janssen, Merck, Novartis, Roche, Sanofi, and Teva; participated on an advisory board for Biogen, Janssen, Merck, Novartis, and Roche; and received support for attending meetings or travel from Biogen, Janssen, Merck, Novartis, Roche, and Sanofi.

DH: none to declare

NV: none to declare

JB: none to declare

KG: none to declare

LB: none to declare

MW: none to declare

JG: none to declare

GG: has received consulting or speaker fees from AbbVie, Aslan, Atara Bio, Biogen, Bristol Myers Squibb–Celgene, GlaxoSmithKline, GW Pharma, Janssen–Actelion, Japanese Tobacco, Jazz Pharmaceuticals, LifNano, Merck & Co, Merck KGaA–EMD Serono, Moderna, Novartis, Sanofi Genzyme, Roche-Genentech, and Teva Pharmaceuticals.

